# Statistical Approach Leveraging Genealogies of Populations with a Founder Effect and Identical by Descent Segments to Identify Rare Variants in Complex Diseases

**DOI:** 10.1101/2025.09.16.25335588

**Authors:** Samir Oubninte, Simon Girard, Claudia Moreau, Michel Maziade, Alexandre Bureau

## Abstract

The missing heritability caused by rare variants (RVs) poses a significant challenge to pre-established statistical methods. Our study aims at detecting RVs using identical-by-descent (IBD) segments as a proxy for recent variants in family data from a population with a founder effect for which genealogy is available—a distinguishing feature of our approach. Inferring IBD segments from genotype array data, which is more accessible than whole genome sequences, enables application to large sample sizes. Our approach involves dividing the genome into fixed-length windows, treating each window as a synthetic genomic region (SG), and then identifying groups of affected individuals sharing a specific IBD segment over an SG by analyzing genotype array data to infer pairwise IBD segments. Data from pairwise IBD segments is then used to identify densely connected haplotypes as IBD clusters via DASH. Lastly, we adapt, implement, and evaluate statistics to test for IBD sharing enrichment among affected individuals within SGs. The null distribution of the genome-wide maximal statistic value is obtained by simulating whole-genome transmission in a genealogy using msprime. For application purposes, Eastern Quebec has been studied as an example of a population with a founder effect. Using the BALSAC database to reconstruct the genealogy of 1,200 subjects across 48 schizophrenia and bipolar disorder multi-generational families led to an 18-generation pedigree with 84% completeness at the 10th generation. The statistic denoted as *S*_msg_ for the “most shared haplotype in an SG” exhibits superior power in detecting causal SGs when compared to the adapted *S*_all_ measure and (with a single causal variant in a region) to GMMAT (Generalized Linear Mixed Model Association Test) applied to IBD clusters. Our analysis of data pertaining to schizophrenia and bipolar disorder reveals no regions that surpass the conventional significance thresholds for harboring rare variants associated with these disorders. Two distinct regions—on chromosomes 5 and 11—stand out due to their maximal *S*_msg_ values. These findings underscore the potential of leveraging genealogical data and IBD segments to uncover rare variants in complex diseases.

## Introduction

Complex diseases, or multifactorial disorders, pose a major public health challenge due to their prevalence, morbidity, and economic burden. They arise from interactions among genetic, environmental, and lifestyle factors. Precision medicine seeks to address this complexity by shifting from “diagnose and cure” to “predict and prevent.” While molecular and statistical genetics have elucidated causal genes for many Mendelian disorders, the genetic basis of complex diseases remains incompletely understood. GWAS have identified numerous common variants, yet these explain only a fraction of heritability, leaving the problem of “missing heritability”. One prominent hypothesis attributes this gap to rare variants (RVs) [Genin, 2019]. Rare variants can exert substantial effects on disease risk [Genin, 2019, López-Cortegano and Caballero, 2019, Schork et al., 2009], making their identification critical for understanding disease mechanisms and improving diagnostics and therapies.

Variants are rare because of recent emergence within the past few generations or by being maintained at low frequencies in the population in absence of strong selective pressures. The identification of rare genetic variants contributing to complex diseases presents a formidable array of challenges. Rarity in the population poses a fundamental hurdle in amassing sufficient data for robust analyses. The intricate web of genetic heterogeneity, complex inheritance patterns, and variable disease outcomes adds layers of complexity to the quest for consistent patterns. Existing statistical methodologies thus often prove inadequate to discern associations with these rare variants. Notwithstanding these formidable challenges, methods focused on rare variants have been developed over recent years such as for familial studies: GESE, rareIBD, and pVAAST, which Bureau et al. compare with their RVS method (for rare variant sharing) [Bureau et al., 2019]. These methods assume genomic sequence is available on the study subjects. Indeed, genomic sequencing is widely regarded as the gold standard for comprehensive variant detection. However, its accessibility is constrained by substantial costs. The approach we propose offers an alternative by relying on genotyping chips rather than genomic sequencing—a common practice in IBD mapping [Chen et al., 2023]—which enables the inclusion of larger sample sizes.

The aim of our methodology is to identify well-defined genomic regions that can be interrogated for potentially pathogenic, rare genetic variants associated with disease. A key contribution of our approach is the integration of genealogical data, which enables us to leverage population structure to enhance the IBD segments reflecting recent haplotype sharing. These IBD segments serve as proxies for recent variants—typically rare and difficult to detect through conventional methods [Gusev et al., 2011]. Leveraging these proxies in conjunction with genealogical data, we endeavor to identify pathogenic genes. These approaches enable the evaluation of co-segregation with the disease, shedding light on the elusive associations that might otherwise remain concealed within the population [Bureau et al., 2014]. The use of genotyping chip data mitigates cost barriers and improves the practicality of genetic studies, albeit within the constraints imposed by the requirement for a genealogy.

In this article, we evaluate power to detect rare variants through IBD segment sharing within relatively isolated populations experiencing rapid demographic expansion, characterized by a distinctive genetic variation distribution typical of populations with a "founder effect". The founder effect should simplify the localization and identification of genes responsible for hereditary diseases compared to the challenges posed by larger and more heterogeneous populations. Typically, affected individuals share a DNA segment inherited from a common ancestor, often one of the population’s founders. The Quebec population exemplifies such a scenario, having been shaped by a major founder effects [Anderson-Trocmé et al., 2023], and is a perfect case for the application of our method. Genealogical data for Quebec is sourced from the BALSAC population database, which contains records spanning from the early European settlement in the 17th century to the contemporary period. These data originating from the digitization of civil records have been interconnected to reconstruct family lineages over nearly 400 years (BALSAC website: https://balsac.uqac.ca/).

Our methodology comprises several steps to identify causal regions, as detailed in the Materials and methods section. We segment the genome into fixed-length windows that are expandable if the IBD structure remains unchanged, and thereafter treat each window as a synthetic genomic region (SG) for association testing using IBD sharing statistics such as our proposed *S*_msg_ statistic. We harness genotyping array data to infer IBD segments serving as the foundation for forming SGs, and then find the groups of affected individuals who share specific IBD segments over SGs, before evaluating statistical methods designed to assess the enrichment of IBD sharing within SGs among affected individuals.

Simulations play a crucial role in this study. First, they provide a null distribution for the genome-wide maximal value of statistics by simulating whole-genome transmission in a genealogy. Through these simulations, researchers can establish a baseline against which observed statistics are compared, enabling the assessment of statistical significance and the identification of shared segments within SG among affected individuals. Second, simulations also intervene in evaluating the performance of the proposed methodology, allowing for the exploration of the method’s robustness across various scenarios and providing insights into its reliability and applicability in real-world settings. We present software implementing the method that exploits the genealogy of a population with a founder effect. This is a feature that most genetic analysis software does not provide [Gauvin et al., 2015].

## Materials and methods

Our approach leverages the genealogy of populations with a founder effect and IBD segments to deduce the genomic regions harboring rare disease risk variants. The methodology follows a structured workflow, as illustrated in Figure 1. Overall, the efficacy of the described strategy depends critically on the selection of accurate and computationally efficiency software tools. All software used in the analysis is documented in the Supplementary Methods. First, we employ the RefinedIBD software package to infer shared IBD segments between pairs of individuals [Browning and Browning, 2013]. Next, these segments are grouped using the DASH software package, which identifies sets of individuals sharing the same IBD regions [Gusev et al., 2011]. Finally, as an original contribution of this work, we introduce the concept of synthetic genomic regions, which harmonize overlapping IBD segments within a region (see subsection Building synthetic genomic regions for more details) to enable statistical testing.

**Figure 1.**
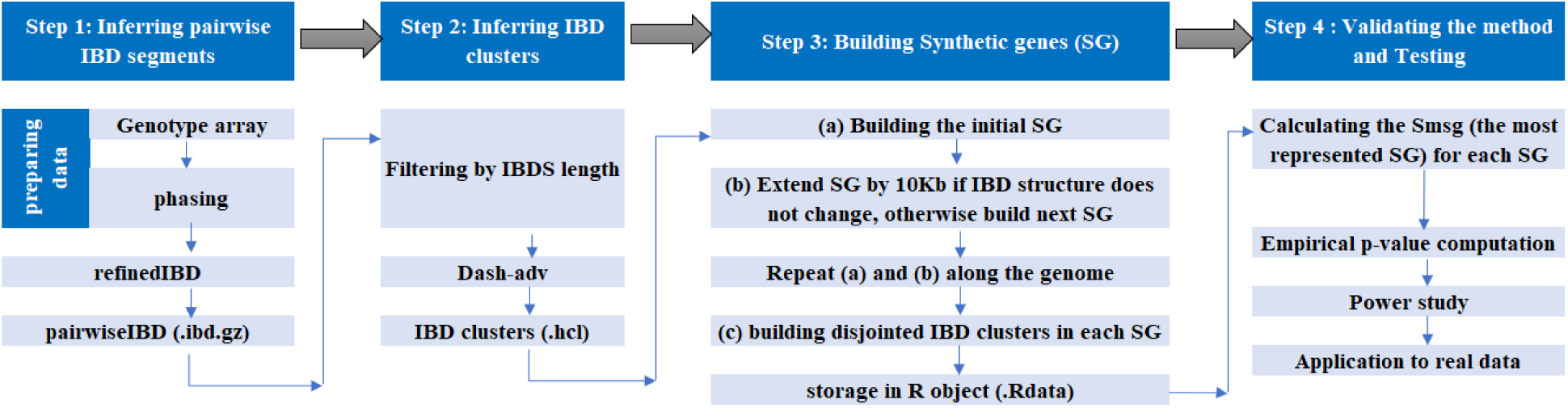
Schematic Overview of the Methodology: This figure illustrates the comprehensive four-step process for analyzing genetic data, including inferring and clustering IBD segments, the innovative construction of synthetic genomic regions, and the validation protocol employed to ensure the method’s accuracy in elucidating genealogical relationships within a population with a founder effect in order to identify causal variants.

We used a family-based case-only design, which reduces potential bias factors associated with unaffected individuals and, moreover, the lack of genetic data from non-affected subjects. To assess whether there is an enrichment of IBD sharing among affected subjects in relation to their genealogical relationships in the population with a founder effect, the *S*_msg_ statistic was developed, and then the method was evaluated.

### Inferring pairwise IBD segments

IBD segments are regions of DNA shared between individuals, inherited from a common ancestor, reflecting the recombination that a chromosome undergoes. Even if genotyping array data usually contain mostly frequent variants, we have kept only variants with minor allele frequency (MAF) greater than 0.1, as recommended by Browning and Browning [2013], prior to haplotype phasing using Beagle [Browning and Browning, 2007]. Phasing is required to generate pairwise IBD segments by RefinedIBD (step 1 of Figure 1). RefinedIBD is a software package that implements a hidden Markov model (HMM) to identify genomic regions that are IBD between two individuals. It also performs genotyping error correction and missing genotype imputation. We applied a log of odds (LOD) score threshold of 3.0 to select IBD segments, as suggested by the authors. It is possible to run Refined IBD several times with different random-number seeds and to merge the resulting IBD segments [Browning and Browning, 2013]. RefinedIBD has an option to merge IBD segments that overlap across different runs, which can improve the detection of long IBD segments. However, this option also has some limitations, such as losing the haplotype information (which is essential for the subsequent analysis) and increasing the running time. The outputs of RefinedIBD allows DASH to infer IBD clusters, as shown at Step 2 of Figure 1 and detailed in the next section.

### Inferring IBD clusters

The second step of the method concerns the construction of IBD clusters (groups of affected individuals sharing the IBD segment). The majority of techniques for creating IBD clusters reported in the literature [Gusev et al., 2011, Moltke et al., 2011, He, 2013, Qian et al., 2014, Purcell et al., 2007] use the IBD segments that are shared by pairs as input. At the beginning, we identified three potential methods: DASH [Gusev et al., 2011], IBD Groupon [He, 2013], and EMI [Qian et al., 2014]. In the absence of implementations of the EMI [Qian et al., 2014] and IBD-groupon [He, 2013] methods, we turned to DASH. DASH scans the genome using sliding windows and generates haplotype subgraphs. It constructs clusters from the largest connected component and iteratively divides them into smaller clusters of desired density (with subgraph density threshold as input parameters) using a minimum cut algorithm (see Gusev et al. [2011] for more detail). One of its two versions (dash-cc) is almost parameter-free. We leverage the more sophisticated and dense subgraph clustering version of DASH (dash-adv), which allows more control over input parameters. The dash-cc version is used in comparative analyses with a competing approach.

### Building synthetic genomic regions

DASH generates a haplotype cluster file, where each line represents an IBD cluster. These clusters exhibit variations in segment lengths, start positions, and end positions. The concept of synthetic genomic region deals with the diverse dimensions of the clusters. These clusters are not disjointed; rather, they exhibit a degree of overlap, reflecting their complex and heterogeneous nature. The SG concept represents segments shared by identified distinct clusters; The process of SG building is simplified and diagrammed in Figure 2 and depicted in more detail in Supplementary Figure S1.

**Figure 2.**
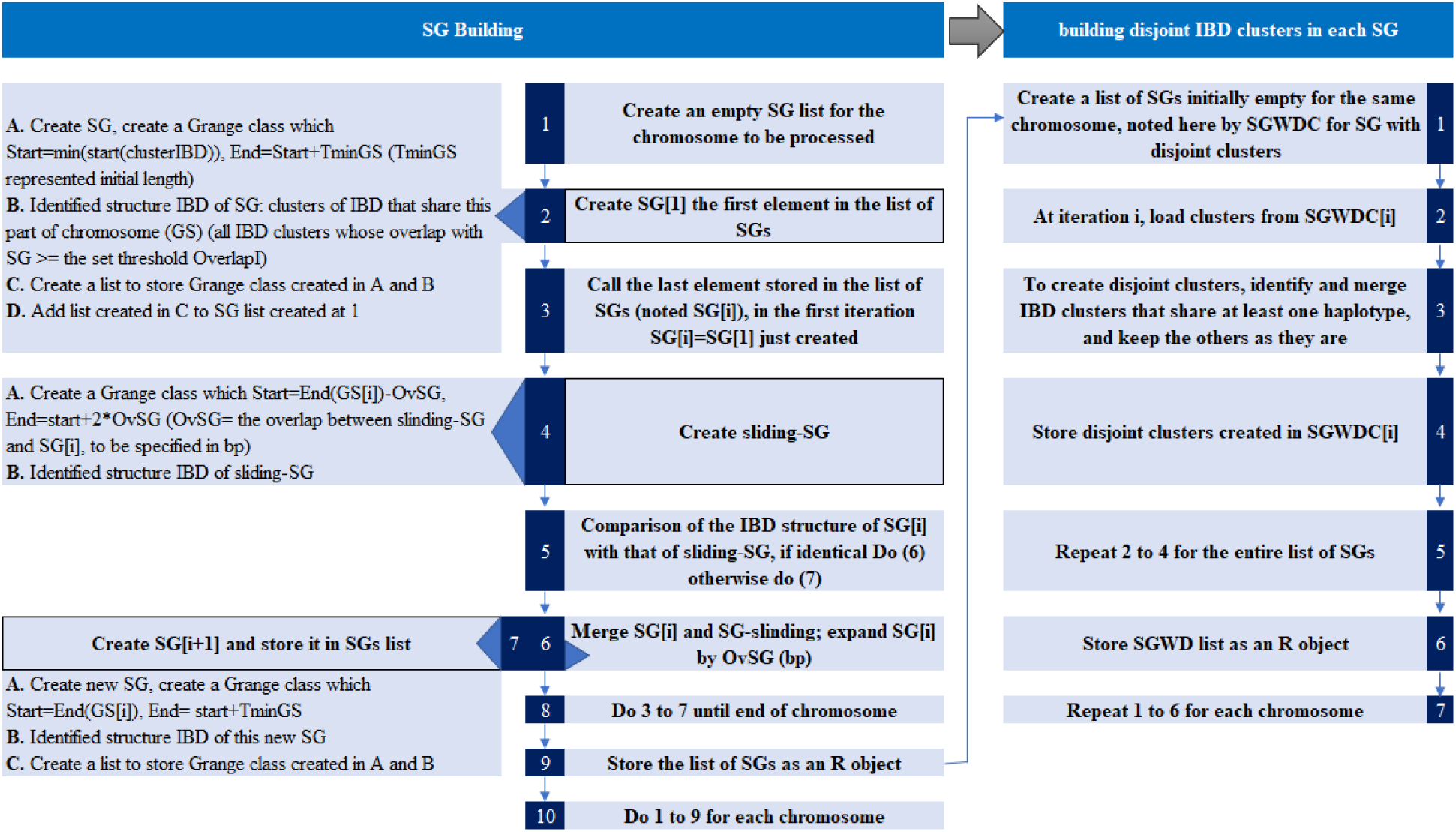
Detailed workflow for constructing synthetic genomic regions and orchestrating IBD Clusters within SGs. This flowchart delineates the algorithmic framework and Bioconductor data structure for generating SGs and handling IBD clusters to ensure accurate genetic analysis.

The process begins by defining an initial SG as a genomic range (Grange). The Grange start position is set to the minimum start coordinate among the identified IBD clusters on a given chromosome, and its width is defined as ***minL*** (Table 1), representing the minimum SG length specified by the user. The IBD structure within the SG is determined by identifying clusters that share the same chromosomal segment and whose overlap with the SG meets or exceeds a predefined threshold, quantified by the parameter overlapI (see Supplementary Figure S1). A similar approach is applied to construct the attaching sliding-SG with a length that must be a fraction of a regular SG. Each creation of an SG is accompanied by the generation of an associated sliding-SG, half of this sliding-SG overlapping with the original SG. If the IBD structures of the SG and its sliding counterpart match, the two are merged; otherwise, a new SG is created, starting at the previous SG’s end and extending by ***minL***. This iterative process - creating SGs and sliding-SGs, comparing IBD structures, merging if required, and storing results —continues until the end of the chromosome is reached. In the second phase, disjoint IBD clusters are established within each SG using an agglomerative approach: the cluster with the most shared haplotype is kept among overlapping ones (see Figure 2).

**Table 1.** Recommended default values for various parameters used in the analysis, along with their descriptions.

| Parameters | Description | Default Value |
| --- | --- | --- |
| minL | Minimum length of SG Grange in Kbp | 400Kbp |
| IBDClusters | DASH output in data frame format | - |
| overlapSS | Overlap between Sliding-SG and SG grange | 10Kbp |
| overlapNS | Overlap between the SG grange and the Next-SG grange | 0 |
| overlapI | Threshold at which the cluster is considered to be included in the IBD structure | 0.5 (50%) |

This procedure ensures that each SG is built with consideration for the overlapping IBD clusters, with a clear method for expanding SGs based on the IBD structure and overlap values, and the data is stored for further genetic analysis. Concretely, an SG is a list object created to store DNA segment information and the clusters of haplotypes sharing this DNA segment. In the subsequent step (Step 4 of Figure 1), these lists are encoded to be tested for elevated IBD sharing.

### Validating the method and testing

This section outlines Step 4 of a process, it is divided into a sequence of tasks: first, calculating the *S*_msg_ (see Equation 1), which stands for the number of copies of the most shared haplotype among affected individuals for each SG identified (i.e., the number of members in the most shared haplotype cluster among cases). *S*_msg_ was inspired by classic sharing statistics such as Smost [Bureau, 2001, Baird et al., 2005] and others [McPeek and Strahs, 1999, Sengul et al., 2001, Basu, 2007, Shugart et al., 2002] that seek to test whether there is an enrichment of IBD segment sharing among affected subjects compared to their genealogical relationships. We will take advantage of the founder effect characterizing the population and the availability of individual genealogies combined with genetic data. In this stage, we will look specifically for IBD segments that are shared more often by patients than what is expected from their genealogical relationships in a Quebec population with a founder effect. To achieve this, we simulate the transmission of the entire genome through generations using a coalescent model constrained by the sample genealogy, which allows us to account for the specific demographic history of the Quebec population. By doing so, we generate a null distribution of IBD sharing under the assumption that no disease-associated variants are present, based on imitated Single Nucleotide Polymorphisms (SNP) arrays (see Simulation study section). Following this, an empirical p-value is computed to determine the significance of the obtained results.

If we have *m_i_* disjoint clusters in *SG_i_*, let *n*_1_, *n*_2_, …, *n_m__i_* represent the number of haplotypes in each cluster (i = 1, …, I = number of SG constructed).

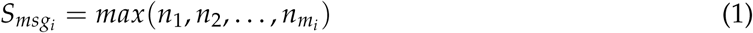

Our genealogy simulations (see Simulation study section) serve two distinct objectives. The first objective is to obtain the null distribution of our statistic, which is calculated on a set of affected subjects. This process solely requires the affected subjects’ genealogy. The second objective is to generate disease scenarios from rare variants. This necessitates both the affected subjects’ genealogy and the genealogy of a reference group. To make the most of the simulated data, we have done both together (see more details in next section). We generated 100 random whole genome sequences (WGS), thereby creating a null distribution for the test statistic. The observed test statistic was then compared to this null distribution to assess its extremeness.

### Study data

In this research, we employed two datasets: one pertaining to schizophrenia and bipolar disorder (SZ-BP), and the other to a Quebec population sample from the CARTaGENE project. For the study of schizophrenia and bipolar disorder, the sample was composed of multigenerational families from the Eastern Quebec population. All subjects were of white race and French-Canadian descent. Personal interviews, family information, and a thorough review of patients medical records (both hospitalized and out-of-hospital) were used to determine the most likely DSM-IV diagnosis over the course of the participants lives. Signed consent was obtained from all participants or from the parents for participants under 18 years of age for collection of all data analyzed here, under the supervision of the University-affiliated neuroscience and mental health ethics committee. The BALSAC database, a comprehensive repository, contains data on the Quebec population from the beginning of European settlement in the 17th century to the contemporary period. Utilizing the BALSAC database, we have meticulously reconstructed the genealogy of approximately 1,200 genotyped individuals across 48 distinct families, inclusive of 400 individuals diagnosed with schizophrenia or bipolar disorder, falling under a broad phenotype. This encapsulates a total of 279 subjects affected by these conditions, conforming to a narrow phenotype (See Supplementary Figure S6a). A cohort of 173 affected probands, defined as individuals without offspring in the genealogy, was subjected to analysis. This analysis cohort excluded two families whose members formed a distinct group on the first two principal components (New Brunswick and Îles-de-la-Madeleine region; Supplementary Figure S6b). The genotyping of the SNP array was conducted in two distinct phases. Detailed information regarding the recruitment process, diagnosis, and the initial wave of SNP array genotyping (48% of the genotyped sample) using the Illumina Omni Express array can be found in Roy et al. [1997], Bureau et al. [2017], Boies et al. [2018]. Further insights into the second wave using the Global Screening Array – Multi-Disease (GSA-MD) and the quality control measures implemented can be found in [Bahda et al., 2023]. The analysis was performed using a composite genotyping array, which is a union of the Illumina Omni Express and GSA-MD arrays. Imputation was conducted for the minimal quantity of residual missing genotypes with Impute5 using the WGS from 1,889 individuals from the CARTaGENE population sample as reference panel. As a result, we successfully retained a total of 573,362 SNPs with MAF > 0.1, distributed across the autosomal chromosomes of the human genome.

The CARTaGENE project [Awadalla et al., 2013] is a population-based biobank cohort study conducted in the Province of Quebec, Canada. The study randomly selected residents aged 40–69 years from provincial health insurance registries to ensure the dataset is representative of the Quebec population. The dataset from this study includes 5,581 probands linked to the BALSAC database.

### Genealogy reconstruction

The reconstruction of the genealogy of the 48 SZ-BP families from the BALSAC database comprises 30,810 individuals (15,355 males and 15,455 females). This genealogy includes 50,646 parent–child links, illustrating the dense network of familial relationships. Its depth is particularly noteworthy, spanning 19 generations between the earliest and latest individuals. Supplementary Figure S7b shows the completeness of the constructed genealogy; mean completeness exceeds 80% at the 10th generation.

To make the most use of the genealogical data and available resources - including time, computing power, and storage - we have combined the SZ-BP genealogy with the CARTaGENE lineage to run extensive simulations. To ensure consistency, we uniformly applied a completeness filtering criterion to the subjects from both the SZ-BP families and CARTaGENE. Our target was to achieve an average completeness for both individuals in a founder couple (as defined in Supplementary Figure S5) of at least 70% by the 10th generation (Supplementary Figure S7a). One founder couple failed to meet this criterion, leading to the exclusion of one entire SZ-BP family.

A total of 6,400 probands satisfied our rigorous selection criteria out of the 8,764 initially considered, including 819 from the SZ-BP families. Connecting the ascending genealogies of these 6,400 individuals yielded a large pedigree comprising 376,026 individuals, with a nearly balanced gender distribution (185,601 men and 190,425 women). Familial connections were defined by 698,585 parent–child relationships. The primary aim of this dataset was to determine the frequency of variants in the population and to pinpoint variants that, while rare in the population, are prevalent in the SZ-BP familial dataset.

### Methods compared

Our novel statistical approach distinguishes itself by capitalizing on the unique properties of genealogies of populations with a founder effect and IBD segments. It harnesses information derived from common variant data on genotyping arrays, more widely available than whole-genome sequencing data. This strategic use of readily accessible data aims to discern rare variants associated with complex diseases. To assess the effectiveness of our method, we conducted a comparative analysis against two well-established approaches: an adapted version of the *S*_all_ measure [Whittemore, 1996] and the Generalized Linear Mixed Model Association Tests (GMMAT) [Chen et al., 2016].

The *S*_all_ measure, introduced by Whittemore and Halpern in 1994 [Whittemore, 1996], has gained prominence for its robustness in detecting genetic linkage. Operating effectively in extended pedigrees, the *S*_all_ measure determines the extent to which alleles at a given locus share a common ancestor. In contrast, GMMAT, a widely-used tool in GWAS studies, specializes in performing association tests. Particularly adept at controlling for population structure and relatedness, the GMMAT is a standard choice in various study designs, making it a pertinent comparator for our methodology. However, it is important to note that, unlike our method, the GMMAT typically requires a control group, which we circumvent by utilizing non-affected individuals as controls in our study. In our implementation, GMMAT was applied to IBD clusters generated by DASH-cc, the recommended version of DASH for association analyses.

Drawing inspiration from Saonli Basu’s [Basu, 2007] work on allele-sharing methods for binary traits in multi-generation pedigrees, we adapted and implemented our version of the *S*_all_ measure. This adaptation proved essential in aligning the method with our specific context, where adapted *S*_all_ measures the degree of haplotype sharing among affected individuals, leveraging the information embedded in the IBD structure of SG.

Consider a vector of length *n*, where *n* is the number of affected, whose *i*-th component is one of the two IBD cluster haplotypes of the *i*-th person at a specific SG. There are 2*^n^* such possible vectors *t*. For each *t*, define:

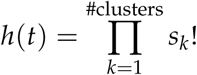

where *s_k_* is the number of haplotypes from IBD cluster *k* that occur in *t*. The score function for a
given SG is:

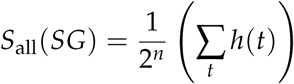

The *S*_all_ statistic assigns greater weight to patterns of haplotype sharing where multiple affected individuals share the same founder haplotype. Unlike *S_msg_*, which considers only the most shared haplotype, *S*_all_ aggregates contributions from several haplotypes.

### Implementation

To translate the theoretical constructs delineated in this manuscript into practical applications, we have created an R package, named FounderRare. This package implements the methodologies expounded in the methods section, including the adapted *S*_msg_ and *S*_all_ segment sharing statistics among other pertinent components. A separate paper [Oubninte et al., 2025] is exclusively dedicated to the FounderRare package and offers an in-depth exploration of its functionalities. The FounderRare package provides researchers the capability to leverage founder effects to identify causal rare variants.

### Simulation study

We used neutral genetic sequence data under a coalescent model simulated using msprime [Baumdicker et al., 2022] to imitate a genotyping array and simulated phenotypes inspired by [Tong and Hernandez, 2020]. Msprime, a coalescent simulation software adept at simulating genetic variation across millions of samples, was selected for its ability to accurately simulate coalescence with recombination for chromosome-sized regions over hundreds of thousands of samples [Baumdicker et al., 2022]. It provides a tree sequence storage file format based on Tskit library, the Tree Sequence Toolkit. Tskit enables efficient storage, manipulation, and analysis of genomes and phylogenies using succinct tree sequences usable in Python, C, and R [Ralph et al., 2020].

Msprime also provides the option to perform discrete-time Wright–Fisher simulations, which are preferable in scenarios where the standard coalescent may be inadequate—such as very large sample sizes or recent migration [Nelson et al., 2020, Baumdicker et al., 2022]. By default, Msprime implements the Hudson coalescent model, a continuous-time approximation of the Wright–Fisher process that is well-suited for simulating genealogical processes with recombination in large populations. The Hudson model introduces mathematical simplifications and focuses on genealogical processes within a known pedigree (https://tskit.dev/msprime/docs/stable/ancestry.html# sec-ancestry-models-specifying).

### WGS simulation

Building simulation models that more accurately capture the genealogical structure[Anderson-Trocmé et al., 2023], we provided to msprime the combined SZ-BP - CARTaGENE lineage reconstructed from the genealogy of the French Canadian population to perform genome-wide coalescent simulations. Msprime traced the ancestry of samples back through the genealogy, accounting for coalescence, recombination, and taking into account relatedness beyond the founders of the known pedigree. We employed an Out-of-Africa (OOA) demographic model [Tennessen et al., 2012] to generate a neutral genetic sequence exclusively for the period preceding the founders of the genealogy. Subsequently, within the genealogical context, we used the fixed pedigree simulation model (see [Anderson-Trocmé et al., 2023] supplementary materials, “Genome simulations” section). Under the OOA demographic model, the European population experienced a series of bottlenecks as they moved out of Africa, into Europe, and eventually to the area which is now the province of Quebec. Using this neutral demographic model, we generate a WGS in Ts files format with a mutation rate of 1 × 10^−8^ and a genetic map chosen to mimic the recombination rate in hg38 [Anderson-Trocmé et al., 2023]. We have generated 100 replicates of WGS for each of the 6,400 probands included in the combined SZ-BP and CARTaGENE genealogy, resulting in a total of 640,000 simulated genomes.

### WGS to SNP chip

To imitate a genotyping array, we downsample the simulated WGS data above to match the average distance between variants of our real genotyping chip and the allele frequency spectrum of the SNP genotypes in the CARTaGENE sample. The variant positions for inclusion in the chip are randomly selected based on probabilities calculated from a histogram of the CARTaGENE MAF distribution and distance parameters based on real chip data (see subsection Study data for more details about the real data) with respect to the first position, the last position, and the number of SNPs in the chip. A Python script was designed to facilitate the selection of SNP positions from WGS data. The algorithm requires predefined parameter values, including a random seed and a minimum distance between SNPs.

The selection process begins with an empty set and iteratively adds positions sampled at random, weighted according to the empirical MAF distribution and continues until the average distance between SNPs exceeds a target distance and until a predefined number of SNPs have been selected. At each step, constraints such as maintaining a minimum inter-marker distance and achieving a target average spacing are enforced. Positions that fail to meet these criteria are discarded, while accepted positions are removed from the candidate pool to prevent reselection. This iterative approach ensures that the final SNP set reflects the desired allele frequency spectrum and spatial distribution, which is essential for designing an effective genotyping array.

### Phenotype simulation

The process of simulating phenotypes involves several steps. First, select a causal region. The desired chromosome is divided into M bins, or regions of 400 kbp. Notably, this length is significantly shorter than the expected length of an IBD segment, which is calculated as 1/(2*n*) Morgans for a common ancestor traced back *n* generations in a large population [Su et al., 2012]. For the Quebec population, *n* is estimated to be between 20 and 25 generations. Subsequently, the MAF distribution was calculated for the simulated WGS of both cohorts: CARTaGENE and the SZ-BP cohort. Within this context, variants that are common in the simulated WGS of SZ-BP probands but rare in CARTaGENE (<1%) are identified. The genomic region maximizing the count of such variants per chromosome is then selected. And then, we code the three possible genotypes for identified variants within the chosen region into a genetic covariate stored in matrix (**X**). The phenotype assignment for individuals under alternative hypotheses is determined by applying an additive model, as shown by equation 2, a logistic regression model based on one or two identified variants sampled from the selected region, depending on the desired situation. We have made an assumption of a disease prevalence of 0.01 when *X* = 0. Given risk variants in our model are rare, the overall disease prevalence is also close to 0.01, representative of the risk of schizophrenia in the population [Bureau et al., 2012].

Let *Y* be the phenotype of interest, a binary disease trait. We consider the logistic regression model:

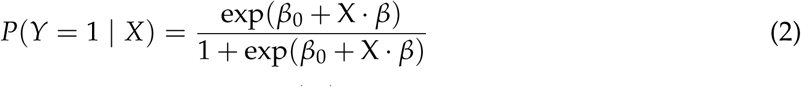

where *β_i_* is the genetic effect of the variant *i*, and exp(*β_i_*) represents the odds ratio comparing *X_ij_* = 1 to *X_ij_* = 0 and *X_ij_* = 2 to *X_ij_* = 1 for individual *j*.

In our pursuit of authenticity and to mirror the recruitment conditions of familial samples, each family is required to include at least two affected individuals, with a minimum total of sixteen affected individuals across all families (e.g., eight families with two affected members each). This design reflects the structure of our real data, which originate from a family study conducted in Eastern Quebec. Populations with founder effects such as this one are particularly conducive to family-based genetic studies, as reduced genetic heterogeneity and elevated frequencies of certain rare variants enhance the power to detect genotype–phenotype associations.

When simulating phenotypes based on a single variant, the variant is selected at random among those common in the simulated WGS of SZ-BP probands and rare in CARTaGENE. In scenarios involving two variants, the selected pair is drawn from different haplotypes within the same causal region. This methodological approach is designed to reflect the complex realities of genetic inheritance, particularly in populations with a founder effect, where extended haplotypes and shared ancestry are common.

## Results

### Validation of generated chips from simulated WGS

We generated SNP chips for 819 probands from simulated WGS data. This validation study was performed on a subset of 178 affected probands. Our aim was to ensure homogeneity of different distributions and concordance between real and simulated chip data, avoiding regions with an apparent excess of IBD in the simulated data. To achieve this, we compared the distribution of IBD segments. In order to align our study with real-world conditions and meet the specific requirements of our research, we selectively retain pairwise IBD segments that are longer than 1.5cM, which we approximated as 1.5Mbp. Additionally, we excluded any segment overlapping with annotated genomic gaps, using the gap list extracted from the Genome Browser annotation track database.

Supplementary Figure S2 presents the length distributions of pairwise IBD segments, demonstrating that, as anticipated, the two distributions are generally similar (see Supplementary Table S1 for more details). The simulated data encompasses a range of values comparable to that of the real data. Furthermore, we observe a marginally higher number of short segments in the real data, which is expected due to genotyping errors inherent in real datasets. Subsequently, we analyzed the quantity of IBD segments that cover specific regions of the genome, as depicted in Supplementary Figure S3. Our findings reaffirm that the distribution of simulated data aligns well with the real data. Next, we concentrate on segments smaller than 10 Mbp to examine the length distribution of haplotype clusters, as illustrated in Supplementary Figure S4. We arrive at the same conclusion. This congruence suggests that the simulated data is replicating the key statistical properties of the real data. Overall, through these analyses, we validate the chips generated from simulated WGS data, thereby confirming their similarity to real data in terms of IBD sharing. This validation provides a robust foundation for downstream analyses.

### Method Validation

To validate our significance assessment procedure, we conducted a series of analyses using both simulated and real SNP chip data. These analyses were performed using the real phenotype and were restricted to affected probands. The validation process involved comparing the distribution of the *S*_msg_ statistic across SGs and chromosomes between the two datasets. As shown in Figure 3, the *S*_msg_ distributions in the real and simulated data show close agreement.

**Figure 3.**
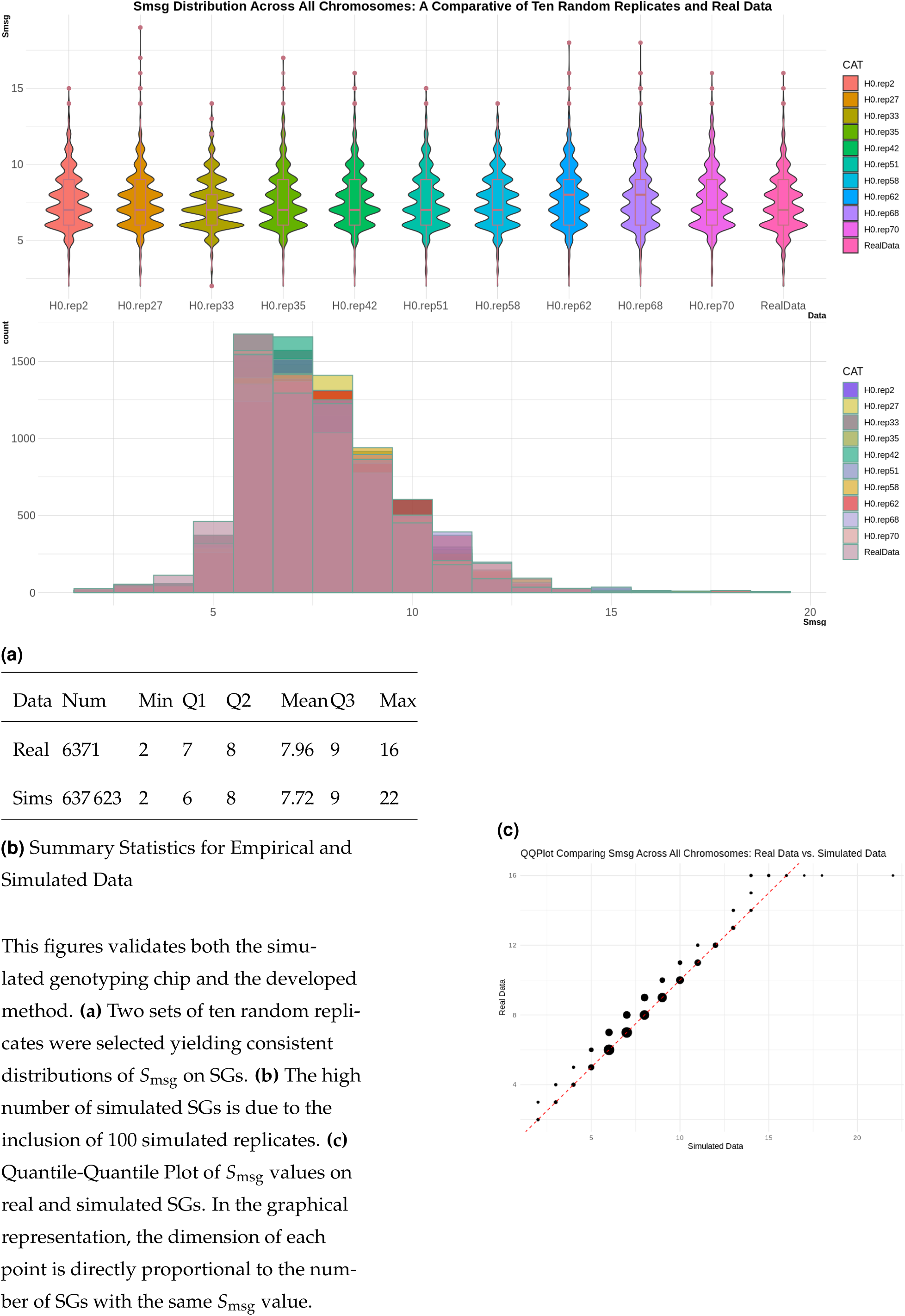
**Comparative Analysis of Smsg Distributions Across Chromosomes**

In Figure 3a, we randomly selected two sets of ten replicates from the simulated data and compared them to the real data. The first set is shown in the violin plot (top), and the second set is presented in the histogram (bottom). The replicates exhibit distributions similar to that observed in the real data. Note that the visible fluctuations ("wiggles") are due to the discrete nature of the *S*_msg_ values, which are integers.

Figure 3b presents the summary statistics of *S*_msg_ for both real data and all simulated replicates, showing close agreement with minor variations in the mean and maximum values. A focused comparison of the summary statistics for the real data and a randomly selected subset of ten simulated replicates (see Supplementary Table S3 for details) further supports this observation. Key statistical measures—including the minimum, first quartile (Q1), median, mean, and third quartile (Q3)—are consistently aligned across both datasets. This consistency is further supported by the QQ plot in Figure 3c, which provides a visual indication that the Type I error is appropriately controlled.

In summary, the validation analyses demonstrate that the simulated data reproduce the behavior of the *S*_msg_ statistic observed in the real data. The alignment of summary statistics, distributional patterns, and visual comparisons supports the reliability of our method.

### Simulated phenotype

In our study, we selected a causal region on each chromosome in every replicate of simulated WGS, for a total of 2200 regions. Under the scenarios involving one causal variant, we obtained 2200 groups of simulated affected subjects meeting recruitment conditions (refer to subsection Phenotype simulation for more context). In scenarios involving two causal variants, the number of affected groups was reduced. This is explained by instances where only one of the two selected variants was present in the simulated data, as shown in Figure 4b. The selected pair must originate from different haplotypes within the same causal region—a condition that is not always satisfied (refer to subsection Phenotype simulation for more details). As anticipated, Figure 4a demonstrates a relation between the number of variants and the number of affected individuals: introducing two variants in the "Two Rare Variants" scenario results in more individuals being affected compared to the "One Rare Variant" scenario.

**Figure 4.**
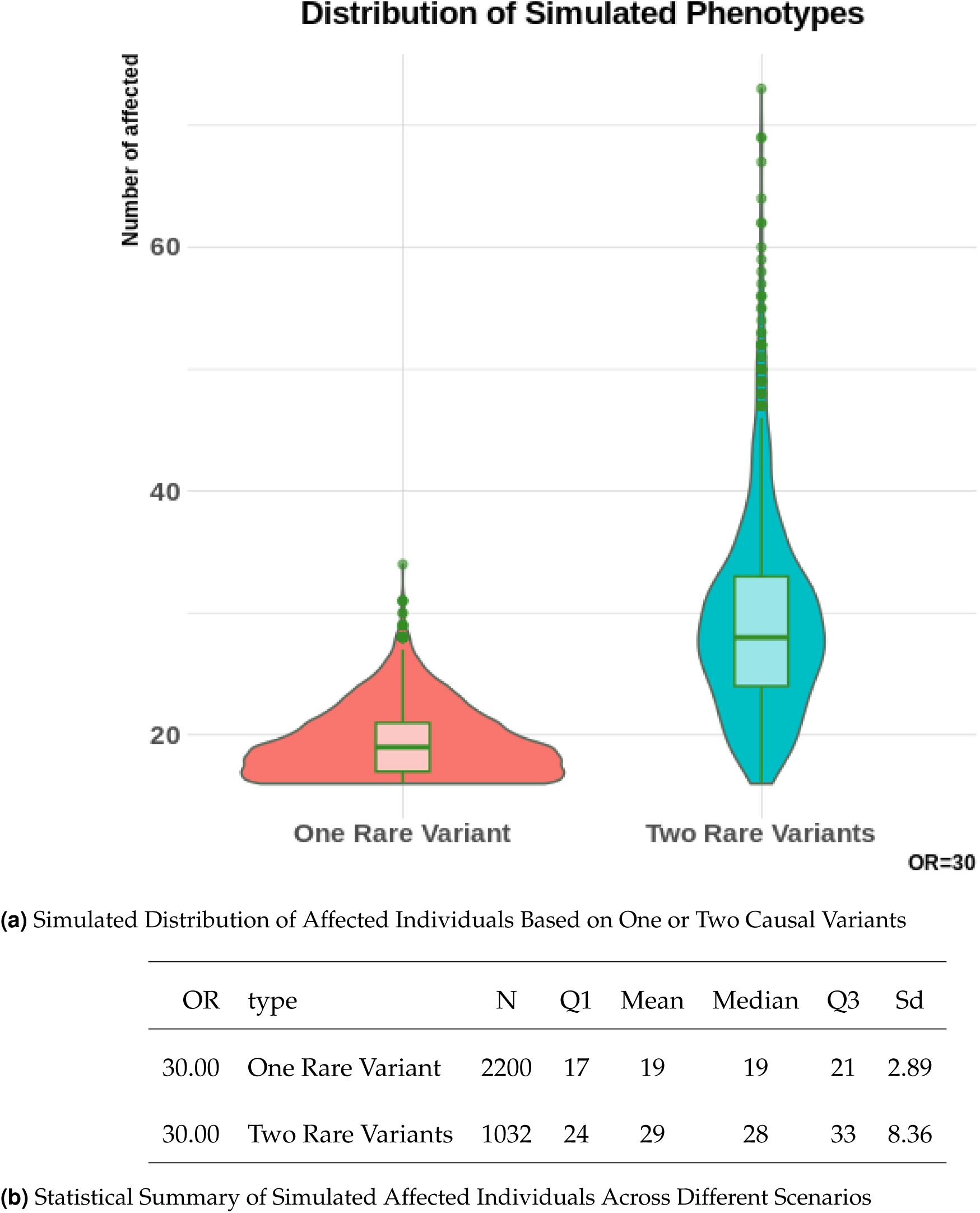
**Statistical Analysis and Distribution of Simulated Affected Individuals Based on One or Two Causal variant; OR: Odd Ratio**

### Power study

To evaluate the effectiveness of our novel statistical approach, we performed a comparison with two recognized methods (Refer to subsection Methods compared): the Adapted *S*_all_ measure and the GMMAT Method. Given the constraints of computing time and memory, the power of the study was gauged using an effect size expressed as an odds ratio of 30 for both the singlecausal-variant and dual-causal-variant scenarios. The power of a test statistic is quantified as a success rate, where success is defined by the identification of a segment overlapping the true simulated causal region as significantly associated with the disease (*S*_msg_ of causal region exceeds a predetermined quantile of the distribution of *S*_msg_ under H0). For *S*_msg_ and *S*_all_, the threshold was set at the 95th percentile of the null distribution of the maximum over the genome. For GMMAT, Bonferroni-adjusted significance thresholds were applied based on the number of tests, using *α* = 0.05/(number of tests).

For the computation of the *S*_msg_ and *S*_all_ statistics under the alternative hypothesis (H1), we utilized all simulated cohorts of affected individuals. In contrast, to assess these statistics under the null hypothesis (H0), we randomly selected nine cohorts of affected individuals from the simulated data to balance computational efficiency and analysis time. These nine cohorts were fixed, we recalculated statistics (excluding the replicate from which the cohort was originally drawn) for each of the 99 replicates other than the one with causal variants (note that each replicate includes 22 variant sets, one per chromosome). In the case of a single variant, the number of individuals in the nine selected cohorts, displays a first quartile (Q1) of 16, with a mean and median of 21, and a third quartile (Q3) of 24. However, in the two-variant scenario, the Q1 is equal to 19, the mean to 30, the median to 29, and the Q3 to 34. These distributions maintain a similarity to their global distribution (Figure 4a).

For GMMAT, we selected non-affected individuals from the same cohorts used for simulating the affected individuals. The ratio was 10 non-affected individuals for every affected proband. Furthermore, GMMAT requires at least one positive semi-definite matrix to model the covariance structure of the random effects. In our analysis, we employed the kinship matrix computed from the genealogy to account for population structure and relatedness.

In Table 2, we summarize the results obtained by each method, considering a genome-wide significance levels of 5%. *S*_msg_ demonstrates superior power in a single variant scenario, surpassing both the *S*_all_ measure and the GMMAT method. However, in a two-variant scenario, while *S*_all_ puts up a better performance, trailing closely behind *S*_msg_, the GMMAT method surpasses the IBD sharing statistics, owing to the higher number of unaffected individuals used as controls than in the one-variant scenario (using 5,3,1 unaffected individuals for each affected individual as controls yields very low power, results not included). Supplementary Table S2 presents the proportion of instances where *S*_msg_ is maximized in the causal and extended causal regions (400kb to each side of the causal region) across chromosomes 7, 8, and 9, in the context of a single variant, to provide a perspective on how far the signal extends from the causal regions.

**Table 2:** Power of Methods under Different Scenarios: The same procedure and identical nine cohorts of affected individuals were employed to calculate the power of *S*_msg_ and *S*_all_. For GM-MAT, Bonferroni-adjusted significance thresholds were determined by the number of tests conducted.

|  | One Variant | Two Variants |
| --- | --- | --- |
| $S_{\text{msg}}$ | 14.5% | 9.3% |
| $S_{\text{all}}$ | 2.1% | 9.0% |
| GMMAT | 8.6% | 14.8% |

### Investigating Schizophrenia and Bipolar Disorder: An Empirical Analysis of Real-World Data

The *S*_msg_ statistic was employed to to detect regions associated with schizophrenia and bipolar disorder combined. We established the significance threshold using 100 replicates of msprime-simulated SNP chip data on the same cohort of affected individuals under the null hypothesis. At the time of this analysis, msprime exclusively provided genotype data of probands, we were restricted to apply our method to a cohort of 178 affected probands to ensure comparability. The Quantile-Quantile plot (Figure 3c) provides initial indications of absence of regions associated with the disease. This observation is further corroborated by the Manhattan plot (Figure 5) representing the *S*_msg_ values for SGs across the 22 autosomal chromosomes, where no region surpasses the significance threshold of *S*_msg_ = 19. The maximal *S*_msg_ value of 16 is achieved in two distinct regions: The first region is located on chromosome 2, starting from position 46578844 bp. This region spans 10 consecutive SGs, covering approximately 4 Mbp. The second region of interest is found on chromosome 10, beginning at position 100608904 bp. This region encompasses 14 consecutive SGs. Sequencing data over these regions could be examined for rare variants on the shared haplotype.

**Figure 5.**
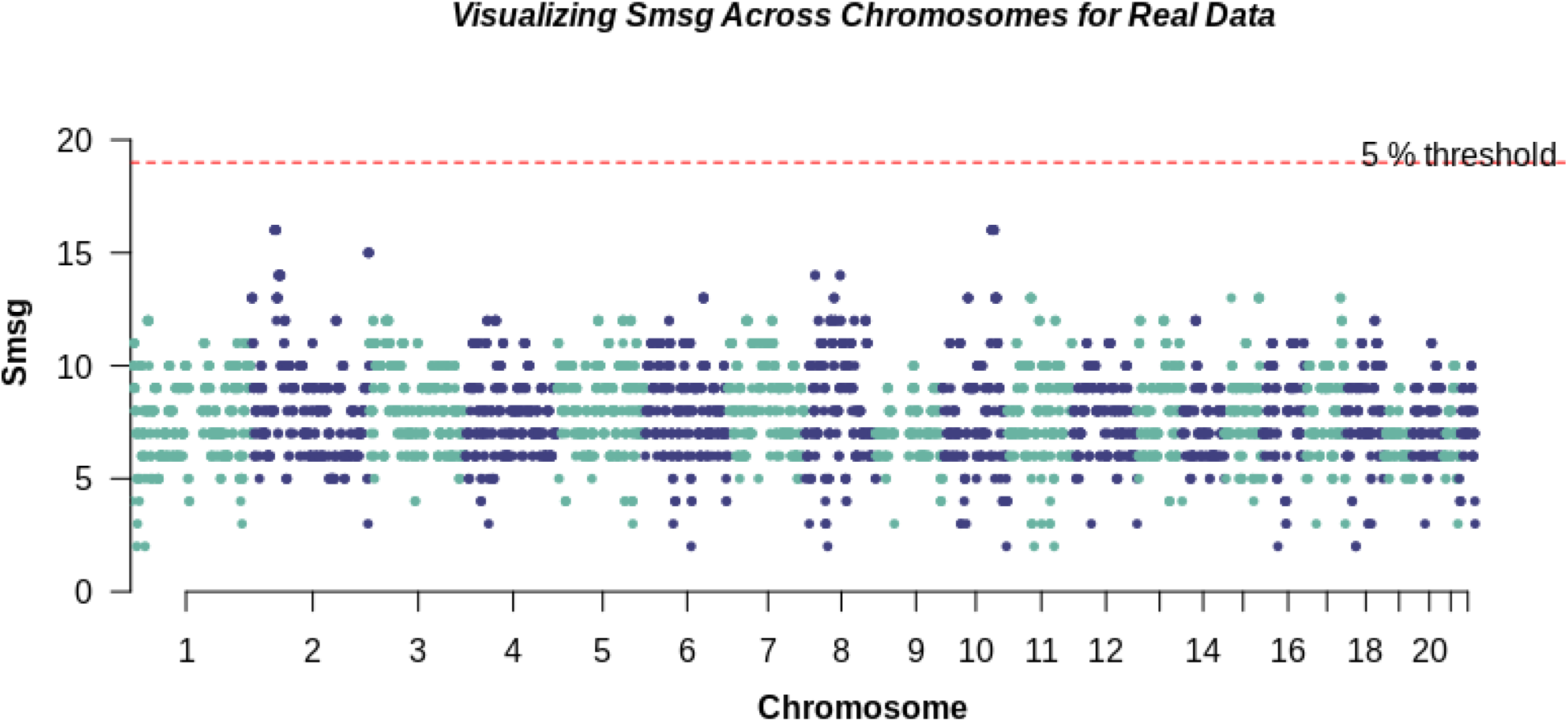
Manhattan Plot Representing the Distribution of *S*_msg_ Across the 22 Autosomal Chromosomes for Schizophrenia and Bipolar disorder Genotype Data (With *S*_msg_ Values on the Y-Axis). The dotted line on the plot represents the 5% genome-wide significance threshold, with points above these lines deemed statistically significant at this levels.

## Discussion

We developed a statistical approach to identify rare variants associated with complex diseases by leveraging genealogical information and identical-by-descent segments in populations exhibiting founder effects. These IBD segments serve as effective proxies for recent rare variants. We specifically focus on pairwise IBD segments within a defined range, typically between 1.5 Mbp and 10 Mbp (see Supplementary Methods). This range covers segments long enough for capturing rare variants in our study population while excluding unexpectedly long segments. Our approach is tailored to populations with founder effects, where a small number of original individuals have contributed disproportionately to the current gene pool. The resulting genetic structure can enhance the power to detect associations that may be missed in more genetically diverse populations [Gagnon et al., 2024].

We developed and evaluated a statistic to test for enrichment of IBD sharing among affected individuals within synthetic genomic regions. The statistic, denoted *S*_msg_, had superior power in detecting SGs containing causal rare variants than the adapted *S*_all_ measure and, in the case of a single causal variant per region, GMMAT (Generalized Linear Mixed Model Association Test) applied to IBD clusters. Notably, *S*_msg_ retains some power even under the stringent condition of a limited number of simulated affected probands, underscoring its potential utility.The observed power remains modest, consistent with previous findings that rare variant association tests often suffer from low power unless causal variants have strong effects or are aggregated effectively. Detection of IBD segments is further complicated by the small size of IBD clusters and background noise from unrelated haplotypes and familial relationships. Moreover, the rarity of the variants themselves limits the number of informative carriers, reducing statistical signal.

In the realm of complex diseases, the role of rare variants cannot be overstated. Although infrequent in the population, these variants can exert profound effects on disease phenotypes, often influencing the onset and progression of complex disorders. Our approach is particularly relevant when, in the context of polygenic architectures, multiple rare causal variants may reside in different genomic regions. Detecting such variants requires sensitive methods, and sensitivity may be improved by accounting for the structure of shared ancestry and haplotype sharing. Our method is designed to tackle this challenge by leveraging the genealogical signal embedded in IBD segments.

Moreover, a strength of our method also appears in its robustness in relation to the choice of parameter values related to IBD structure and density of clusters (results not provided), and unlike GMMAT, IBD sharing statistics do not necessitate a control sample. Furthermore, our method has additional features that allows one to save the constructed SGs with their IBD structure, which can be coded as input to infer other statistics. However, like all techniques, it has limitations. It necessitates a genealogy of sufficient completeness to simulate the sharing of recent IBD segments, and a substantial number of replicates need to be simulated. This may not always be feasible or straightforward in terms of computing resource consumption and speed.

Indeed, we have not yet conducted an exhaustive power study, primarily due to the computational demands of analyzing several whole-genome sequences. Such an analysis would provide a more comprehensive understanding of our method’s ability to detect rare variants across a range of scenarios and in larger samples of affected individuals. The proposed method may have limited power to detect associations with rare variants that have small effects or that affect specific patient subgroups. Additionally, the current implementation may not fully capture the joint effects of rare variants with common variants and environmental factors. Approaches that account for gene–gene and gene–environment interactions will be crucial for unraveling the complex etiology of these disorders. In this context, a novel approach specifically designed for biobank-scale data has emerged that leverages the ancestral recombination graph (ARG) within a linear mixed model (LMM) to identify potentially associated, yet unobserved, genomic variants [Zhang et al., 2023]. This method focuses on continuous phenotypes rather than complex diseases. It is worth noting that its power analyses were conducted on specific genomic regions rather than genome-wide, and primarily involved unrelated samples of homogeneous ancestry. Our method is also applicable to such contexts; however, in this study, we applied it within a familial setting. Future work will involve a comparative study between this method and ours, taking these unique features into account. In particular, we aim to include all subjects with genotype data (not only probands) by developing the necessary extensions.

## Data and code availability

### Data Availability

Genealogical data pertaining to the Quebec population with a founder effect and CARTaGENE can be procured through a request process via the BALSAC platform. This data is provided in an deidentified format to uphold the privacy and confidentiality of the individuals involved. You can find more information and submit your request at https://balsac.uqac.ca/acces-donnees/. The empirical and simulated datasets, integral to the findings of this study, can be made accessible upon formal request.

### Code Availability

Code to run FounderRare is available on GitHub: https://github.com/oubninte/ run_FounderRare. Information about the FounderRare package, and its installation, is available on https://github.com/oubninte/FounderRare. The code used for analyses, visualizations, running genome simulations, extracting rare variants, and simulating phenotypes is freely available on demand.

#### Languages Used

The code encompasses R, Python, and Bash.

## Acknowledgments

We would like to express our heartfelt gratitude to the following individuals and organizations for their invaluable support and contributions to this research:

- Dr Simon Gravel for the invaluable critical feedback and engaging discussions that improved the quality of this work.
- Our dedicated team members, especially Mr. Jasmin Ricard, whose tireless efforts and collaboration made this project possible.
- We are grateful to all the participants who enabled this study by contributing their DNA and to the participants who provided family information enabling the reconstruction of their genealogy. For data from Quebec, we thank the BALSAC team for its management and curation of the genealogy database and the CARTaGENE team and Genome Quebec for their management and curation of genotype data.

