## Supplementary information for "Statistical Approach Leveraging Genealogies of Populations with a Founder Effect and Identical by Descent Segments to Identify Rare Variants in Complex Diseases"

### Supplementary Methods:

The field of IBD segment detection has developed rapidly in the early 2010s in response to the increased density of genetic data along chromosomes, a much higher resolution of IBD segment detection in these data, and the diverse applications of IBD segments (Browning & Browning 2012; Durand et al. 2014; Zuk et al. 2012). Detecting the presence and distribution of IBD segments is fundamental to many genetic applications such as haplotype phasing, identifying disease-associated genes, and estimating the heritability of traits and diseases (Gusev et al. 2011; Browning & Browning 2012; Durand et al. 2014; Zuk et al. 2012).

In the literature, we can find many tools to infer pairwise IBD segments, but each one is more adaptable to a specific need. RaPID (Naseri et al. 2019) stands out for its speed, making it particularly suitable for large-scale biobank datasets. Despite outperforming existing methods in terms of effectiveness and efficiency, especially when it comes to identifying lengthy IBD segments (>5 cM) in simulated data, RaPID approaches tend to overestimate the length of IBD segments by approximately 0.4cM. Durand et al. (2014) addressed the issue of false positives in IBD segments inferred by GERMLINE. Their proposed scoring method (HaploScore) aims to reduce false positives caused by genotyping errors and switch errors.

In their study, Bjelland et al. (2017) evaluated GERMLINE, RefinedIBD, HaploScore, and their FISHER method using both real and simulated genomic sequence data. The study concluded that FISHR is slightly more accurate than all other tools in detecting long IBD segments (>3 cM) but slightly less accurate than RefinedIBD in detecting short IBD segments (<3 cM). This leads us to favor refined IBD methods, as we aim to identify relatively recent rare variants, with IBD segments ranging from 1.5 to 10 cM serving as proxies.

In the context of a population with a founder effect, we are particularly interested in IBD segments that fall within a specific range, typically between 1.5 cM and 10 cM. This range is optimal because it allows us to capture not very long IBD segments, which are likely to capture only family relationships and thus less informative for our purpose, and not very short ones, which may be too common in the population to provide meaningful information about specific genetic variants. By focusing on this range, we can increase the likelihood of identifying rare variants that have a significant impact on the traits of interest in our study population. This approach is particularly powerful in populations with a founder effect, where a small number of original individuals contribute significantly to the gene pool of the current population. The unique genetic structure of these populations can often allow us to detect associations that might be missed in more diverse populations.

Most methodologies for constructing Identical by Descent (IBD) clusters, as documented in the literature, utilize shared IBD segments between pairs as their primary input. Similar to DASH, IBD-Groupon also detects groups of IBD segments based on pairwise IBD segments. It uses a hidden Markov model (HMM) to automatically determine cliques and IBD-group length. Compared to DASH, IBD-Groupon has a similar execution time but higher accuracy for a small sample size of 90 related individuals (He 2013). IBD-Groupon's performance, evaluated on simulated data of chromosome 22 with 6159 SNPs, demonstrates high power in detecting short IBD segments (He 2013).

EMI (Qian et al. 2014) is an algorithm for searching multi-IBD clusters along sliding windows in the genome. It constructs highly connected clusters. Unlike DASH, EMI adopts an agglomerative approach. It constructs each cluster from initial haplotypes and recursively adds qualifying haplotypes to extend the cluster. EMI ensures high computational efficiency through the use of priority queues. Simulations show that the difference in execution time between EMI and DASH becomes more significant in regions with higher pairwise IBD sharing (isolated populations). The relative performance of DASH and IBD-Groupon has not been evaluated in populations with a founder effect, larger sample sizes, or multiple parameter settings. DASH performance in terms of speed and accuracy is comparable to that of EMI (Qian et al. 2014), whereas IBD-Groupon requires a very large memory (He 2013). The unavailability of implementations for both EMI and IBD-Groupon methods made the choice of DASH obvious.

In the field of coalescent simulators, msprime has distinguished itself as a tool of exceptional efficiency. It surpasses other simulators such as MaCS (Chen et al. 2009) , scrm (Staab et al. 2015) , and msms (Ewing & Hermisson 2010) in terms of speed. The design of msprime is tailored to facilitate realistic genome simulation across extensive sample sizes. It employs sparse trees and coalescence records as the primary units for genealogical analysis. This approach enables the exact simulation of the coalescent with recombination for chromosome-sized regions across hundreds of thousands of samples. When pitted against msms, the only other exact simulator compared by (Baumdicker et al. 2022) msprime exhibits significant speed advantages. This efficiency gain is particularly noteworthy considering that msms was excluded from some comparisons due to its slow speed and unreliability for large simulations. Given these attributes, msprime stands as the recommended tool for simulating genetic variation across large sample sizes and long sequences.

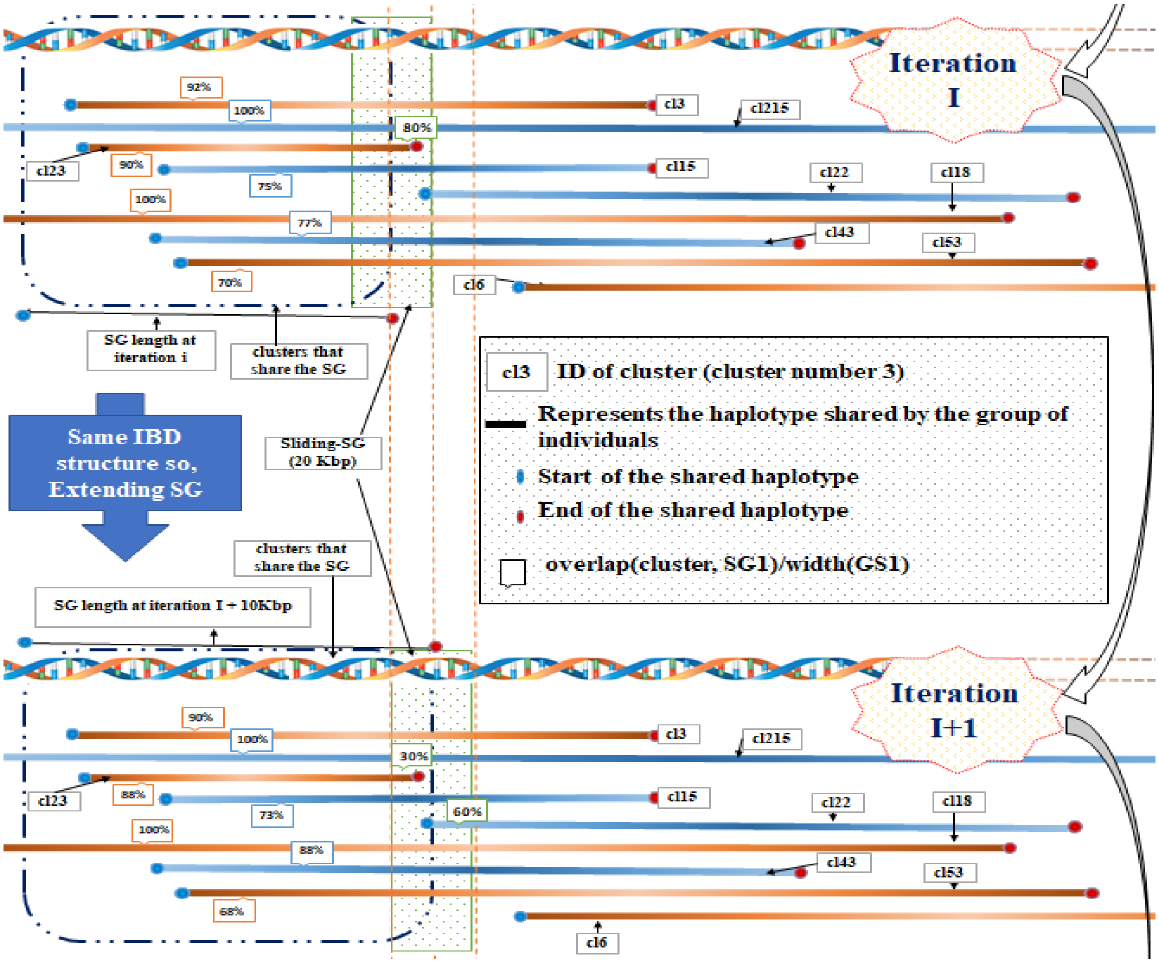

**Supplementary Figure S1:** Schematic representation of the process for identifying and constructing SGs. The figure consists of two parts illustrating two successive iterations (Iteration I and Iteration I+1) of the process. Horizontal lines (blue and brown) represent haplotypes shared by clusters of individuals labeled with IDs such as cl3, cl15, cl22, etc. The lengths of these lines vary, reflecting the diversity of shared haplotypes. These lines are connected by a dashed black line, representing the SG. The percentages along the shared haplotypes indicate the proportion of haplotype overlapping SG or the sliding-SG (the green box). At the bottom of the diagram, a measure of SG length is shown, suggesting that it can change over iterations. In this example, a vertical brown dashed line indicates an increased SG length of "10Kbp" from iteration I to I+1 due to the similarity between the IBD structure of the SG and the sliding-SG (same clusters with >= 50% overlap with the SG also have >= 50% overlap with the sliding-SG). In the forthcoming iteration, it is anticipated that the algorithm will construct a new SG.

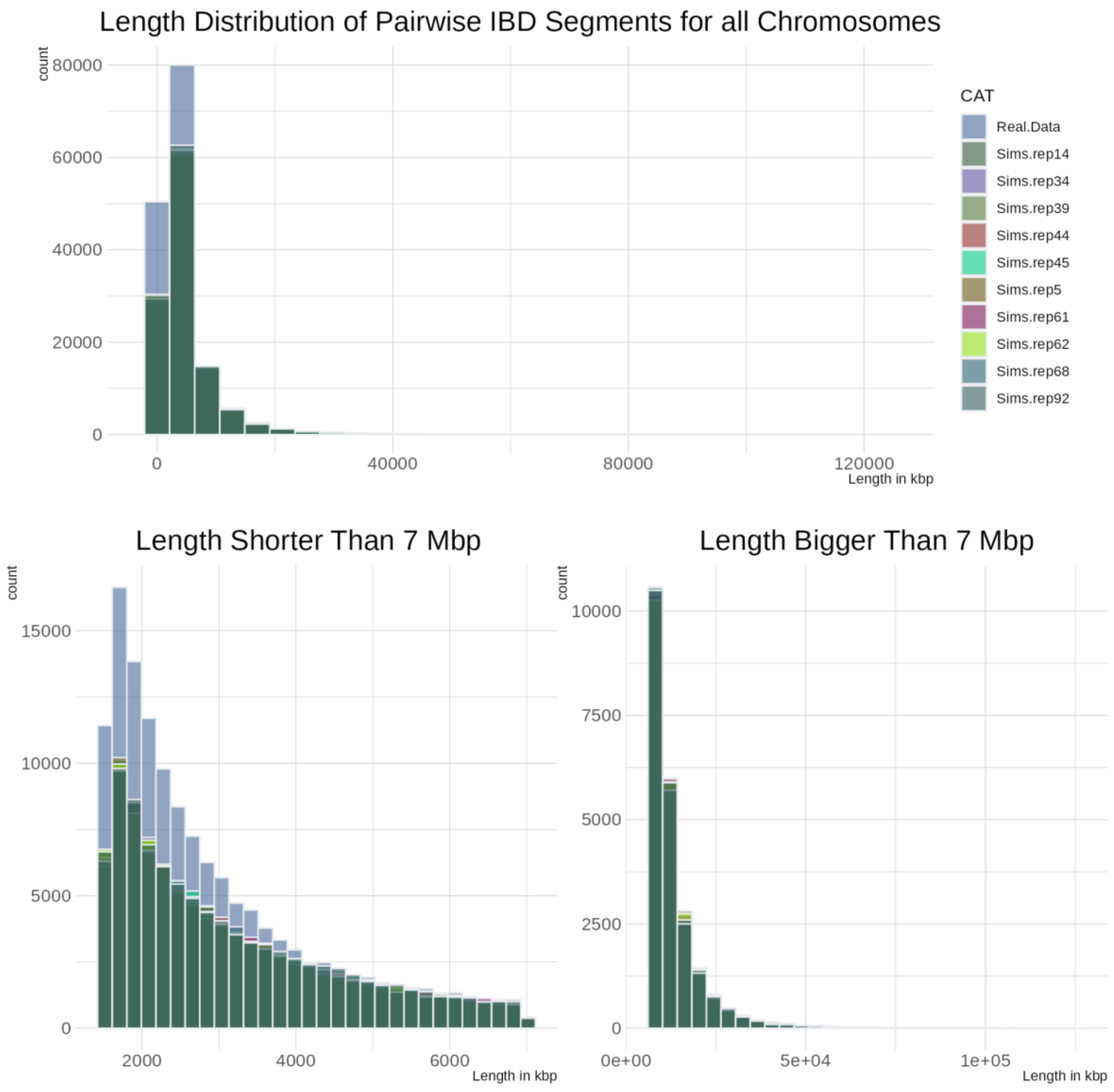

**Supplementary Figure S2:** Presents the length distribution of pairwise IBD segments for real data (based on 178 affected probands) and for ten randomly selected replicates for the same set of probands, across all chromosomes. As anticipated, the two distributions are generally similar. The simulated data encompasses a range of values comparable to that of the real data. However, we observe a higher number of short segments in the real data (sub-figure in the bottom left).

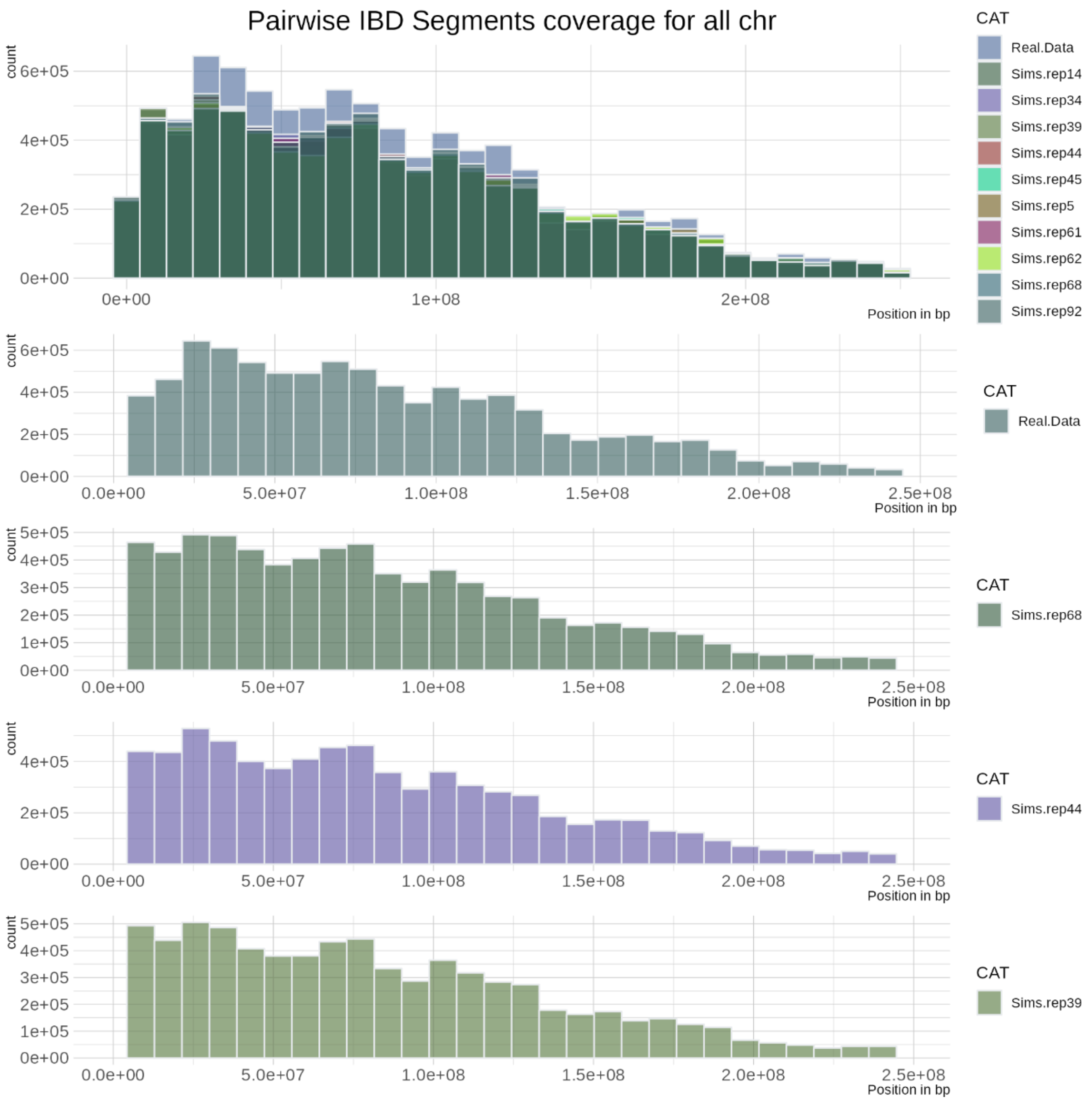

**Supplementary Figure S3:** The quantity of IBD segments that cover specific regions of the genome for real data and ten randomly selected replicates across all chromosomes. Our findings reaffirm that the distribution of simulated data aligns well with the real data.

| DATA | N | Min | Q1 | Median | Mean | Q3 | Max |
|---|---|---|---|---|---|---|---|
| **Real.Data** | 146 404 | 1 500 | 1 916 | 2 597 | 3 671 | 4 052 | 92 718 |
| **Sims.Data** | 11 607 706 | 1 500 | 2 111 | 3 231 | 5 061 | 5 786 | 136 280 |

**(a)** Comparative Analysis of PIBDS Length in Kbp for Real and Simulated Data. The simulated data encompasses 100 replicates.

| CAT | N | Min | Q1 | Median | Mean | Q3 | Max |
|---|---|---|---|---|---|---|---|
| **Real.Data** | 146404 | 1500 | 1916 | 2597 | 3671 | 4052 | 92718 |
| **Sims.rep14** | 115056 | 1500 | 2106 | 3205 | 5044 | 5730 | 106215 |
| **Sims.rep34** | 116030 | 1500 | 2109 | 3257 | 5097 | 5861 | 124765 |
| **Sims.rep39** | 115914 | 1500 | 2100 | 3221 | 5084 | 5767 | 114887 |
| **Sims.rep44** | 115517 | 1500 | 2125 | 3271 | 5113 | 5888 | 103791 |
| **Sims.rep45** | 116600 | 1500 | 2104 | 3202 | 5000 | 5695 | 112635 |
| **Sims.rep5** | 116341 | 1500 | 2091 | 3217 | 5048 | 5809 | 111187 |
| **Sims.rep61** | 115012 | 1500 | 2126 | 3292 | 5118 | 5897 | 102808 |
| **Sims.rep62** | 116487 | 1500 | 2100 | 3224 | 5057 | 5787 | 121009 |
| **Sims.rep68** | 116305 | 1500 | 2116 | 3224 | 5033 | 5780 | 95821 |
| **Sims.rep92** | 118417 | 1500 | 2108 | 3255 | 5018 | 5729 | 97520 |

**(b)** Descriptive Statistics of PIBDS Length in Kbp for Real Data and Ten Random Replicates from Simulated Data.

**Supplementary Table S1**: **Comparative Descriptive Statistics of PIBDS Length in Kbp for Real and Simulated Data.** The table enumerates the number of PIBDS (N), minimum (Min), first quartile (Q1), median, mean, third quartile (Q3), and maximum (Max) length for both Real and Simulated data.

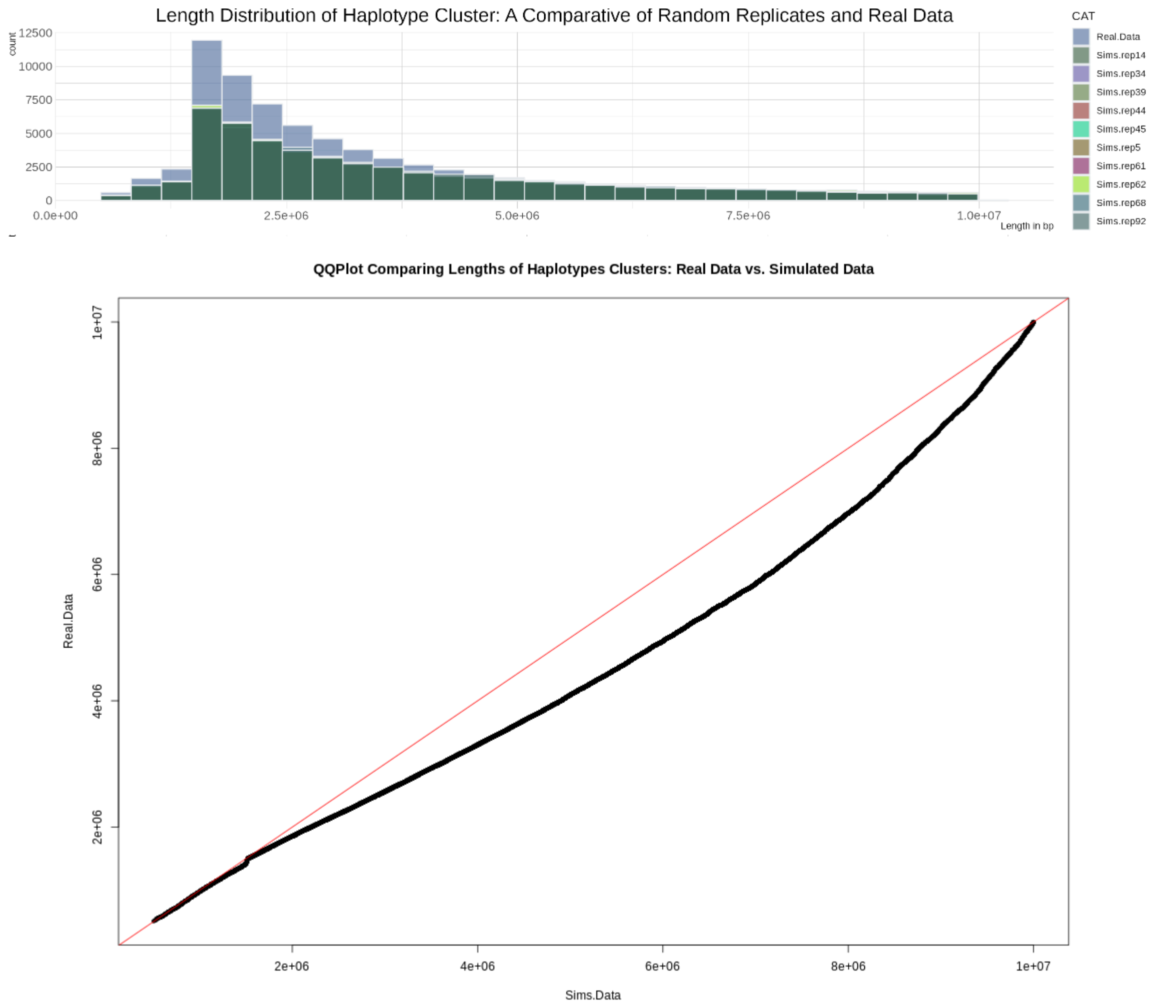

**Supplementary Figure S4:** Presents the length distribution of haplotype clusters inferred by Dash-adv for real data and ten randomly selected replicates, demonstrating that, as wanted, the two distributions are generally similar. The simulated data encompasses a range of values that is comparable to that of the real data. The analysis concentrates on segments smaller than 10Mbp.

| | **Chr=7** | **Chr=8** | **Chr=9** |
|---|---|---|---|
| **causal region*** | 21.5% | 19.4% | 21.5% |
| **Extended causal region*** | 24.2% | 20.0% | 22.6% |

| (+400kb to each side) |
|---|

**Supplementary Table S2:** Comparative Analysis of Maximum Smsg Correspondence to Causal and Extended Causal Regions Across Chromosomes 7, 8, and 9 in case of one variant: This table presents the proportion of instances where the maximum Smsg in the genome corresponds to the Smsg of the causal and extended causal regions.

**Distinct clustering methods: cluster with the most haplotypes are keept among overlapping clusters.*

| Data | N | Min | Q1 | Median | Mean | Q3 | Max |
|---|---|---|---|---|---|---|---|
| **H0.rep2** | 6375 | 2 | 6 | 7 | 7.63 | 9 | 15 |
| **H0.rep27** | 6386 | 2 | 6 | 7 | 7.63 | 9 | 19 |
| **H0.rep33** | 6361 | 2 | 6 | 7 | 7.41 | 8 | 14 |
| **H0.rep35** | 6370 | 2 | 6 | 7 | 7.65 | 9 | 17 |
| **H0.rep42** | 6379 | 2 | 6 | 7 | 7.77 | 9 | 16 |
| **H0.rep51** | 6379 | 2 | 6 | 7 | 7.67 | 9 | 15 |
| **H0.rep58** | 6380 | 2 | 6 | 7 | 7.68 | 9 | 14 |
| **H0.rep62** | 6373 | 2 | 6 | 8 | 7.89 | 9 | 18 |
| **H0.rep68** | 6392 | 2 | 6 | 8 | 7.85 | 9 | 18 |
| **H0.rep70** | 6374 | 2 | 6 | 7 | 7.68 | 9 | 16 |
| **Real.Data** | 6371 | 2 | 6 | 7 | 7.44 | 9 | 16 |

**Supplementary Table S3: Comparative Summary Statistics for Empirical and Simulated Data**. This table presents the summary statistics of the Smsg distribution over synthetic genes across all chromosomes for both empirical and simulated data. The data includes ten randomly selected replicates from the simulation data, demonstrating remarkable similarities with the empirical data. Minor variations in the mean, median and maximum values highlight the robustness of the simulation process.

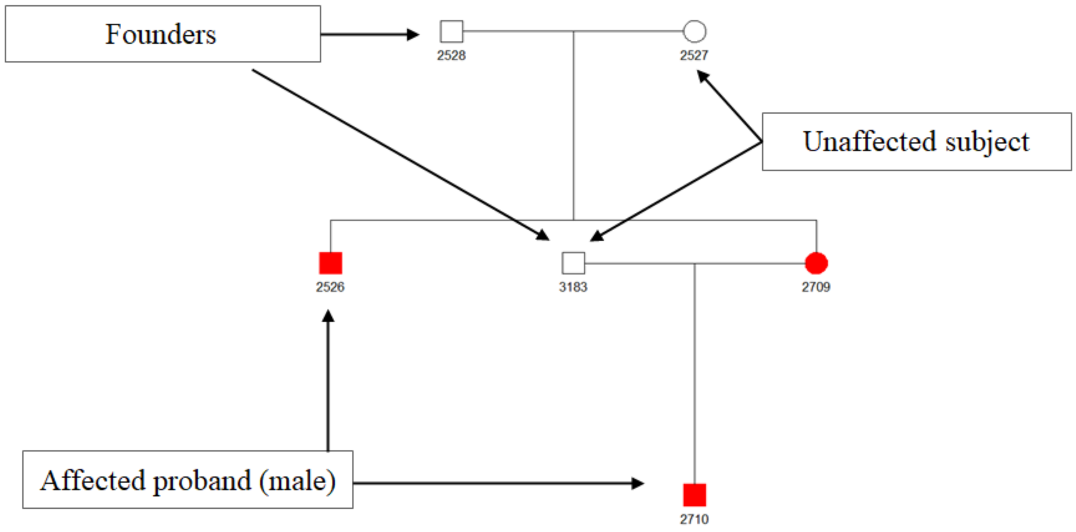

**Supplementary Figure S5:** Example of pedigree structures considered in this study. Family "108" comprises 6 subjects, including 2 probands (individuals without children). Males are represented by squares, females by circles, and affected individuals are highlighted in red. Each individual is assigned a unique numerical identifier. "Founders" are individuals without recorded parents. We removed 2 subjects from generation 2 to prevent identification of the family or its members.

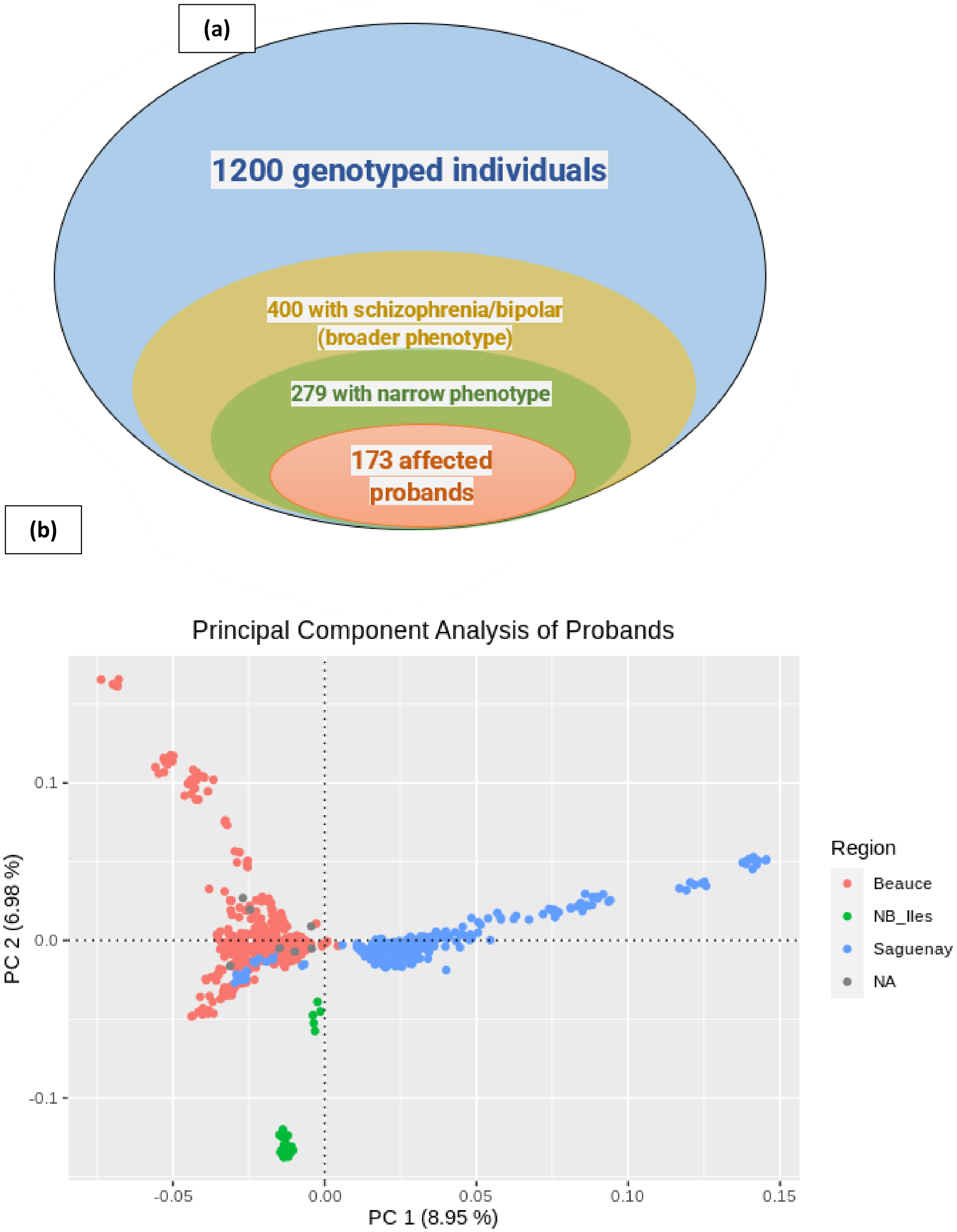

**Supplementary Figure S6 : (a)** Our research encompasses data from 1,200 genotyped individuals spanning 48 distinct families. Within this cohort, 400 individuals exhibit a broad phenotype indicative of either schizophrenia or bipolar disorder. Notably, 279

subjects are characterized by a narrow phenotype. Among these, 173 are identified as affected probands—individuals without offspring in the genealogical records and outside the New Brunswick and Îles-de-la-Madeleine region. **(b)** This Principal Component Analysis (PCA) plot provides a graphical representation of the genetic diversity among children. Each point corresponds to an individual, colored by their region. The axes represent the first two principal components (PCs), which capture the largest variations in the data. The proportion of variance explained by each PC is indicated in the axis labels. NB_Iles: New Brunswick and Îles-de-la-Madeleine.

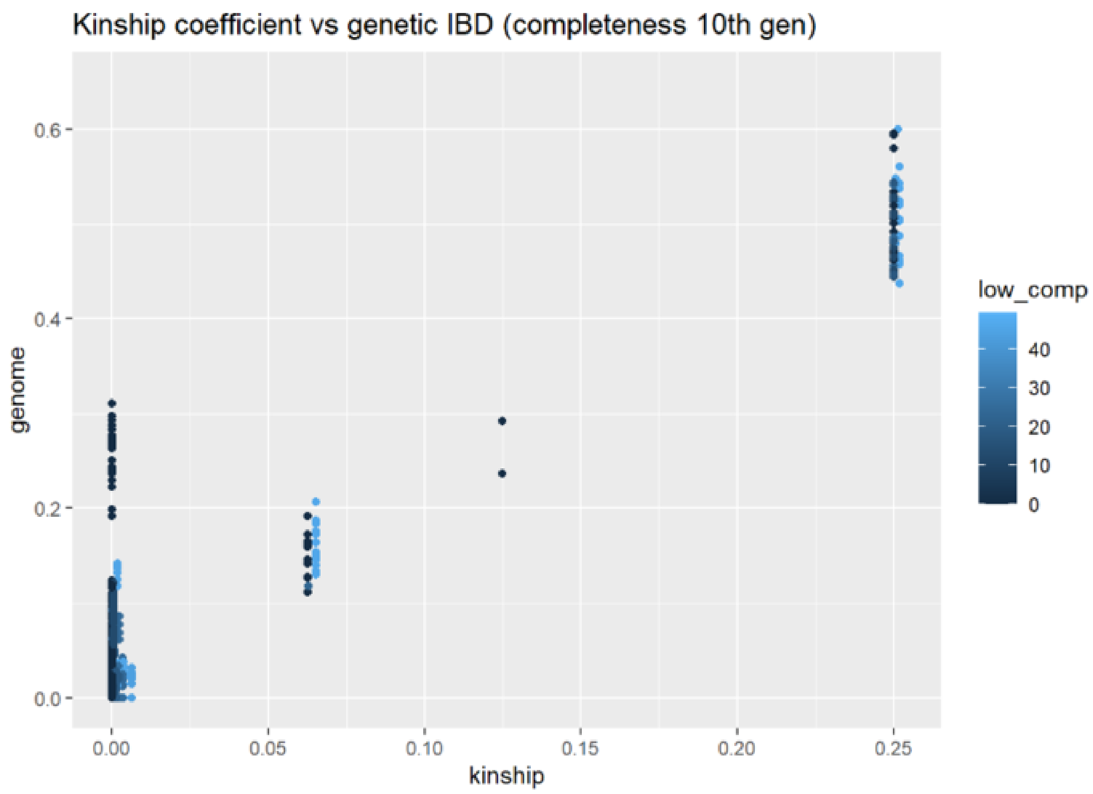

Kinship coefficient vs genetic IBD (completeness 10th gen)
genome
kinship
low_comp
0.0
0.2
0.4
0.6
0.00
0.05
0.10
0.15
0.20
0.25
0
10
20
30
40

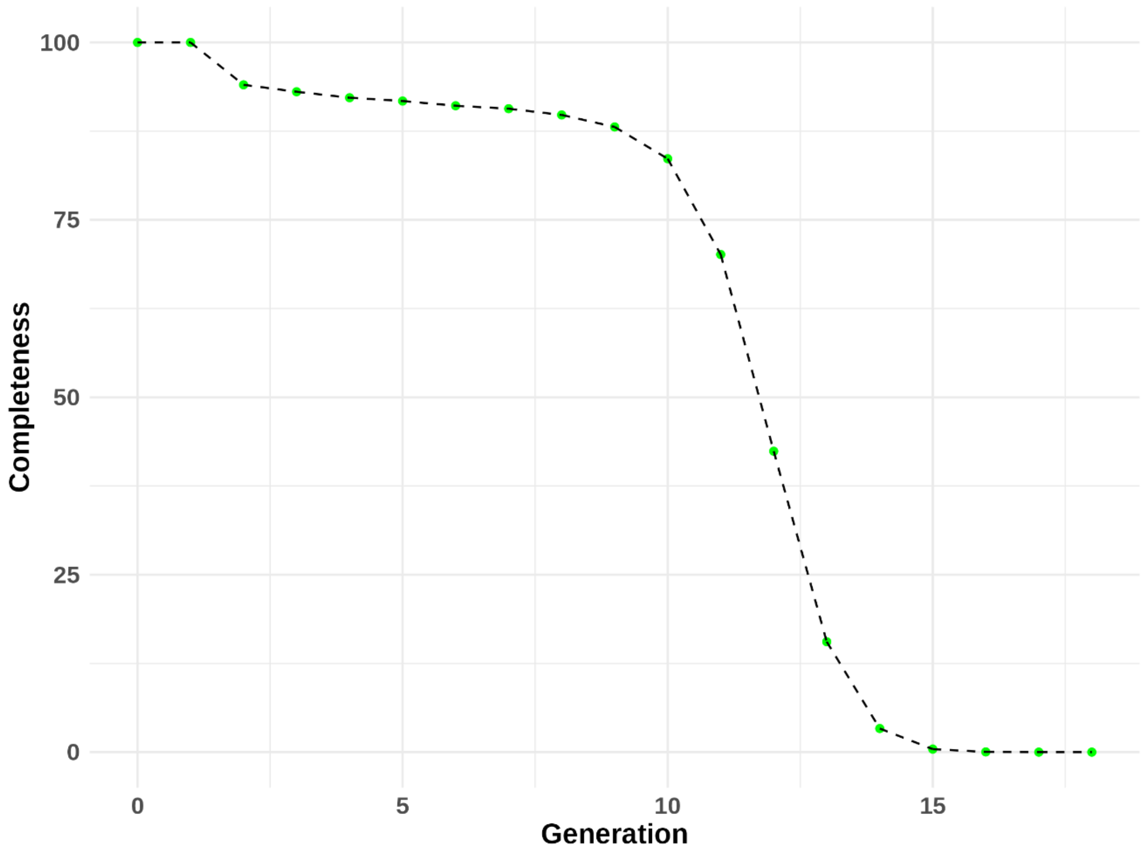

Completeness
Generation
0
25
50
75
100
0
5
10
15

**Supplementary Figure S7 : (a)** Concordance of genealogical kinship with IBD Proportion IBD (i.e. P(IBD=2) + 0.5*P(IBD=1)) for the most recent 10 generations of the genealogy. The color represents the lowest completeness within a pair. It shows very good agreement between kinship and the proportion of IBD shared, except for a small group whose completeness is very low, hence the 70% filter criterion. **(b)** shows the very good completeness of the SZ-BP genealogy that we had constructed thanks to the BALSAC database.
